# Decay of Fc-dependent antibody functions after mild to moderate COVID-19

**DOI:** 10.1101/2020.12.13.20248143

**Authors:** Wen Shi Lee, Kevin John Selva, Samantha K. Davis, Bruce D. Wines, Arnold Reynaldi, Robyn Esterbauer, Hannah G. Kelly, Ebene R. Haycroft, Hyon-Xhi Tan, Jennifer A. Juno, Adam K. Wheatley, P. Mark Hogarth, Deborah Cromer, Miles P. Davenport, Amy W. Chung, Stephen J. Kent

**Affiliations:** Department of Microbiology and Immunology, University of Melbourne, at the Peter Doherty institute for Infection and Immunity, Melbourne, VIC, Australia; Immune Therapies Group, Burnet Institute, Melbourne, VIC, Australia; Department of Clinical Pathology, University of Melbourne, Melbourne, VIC, Australia; Department of Immunology and Pathology, Monash University, Melbourne, VIC, Australia; Kirby Institute, University of New South Wales, Kensington, NSW, Australia; Australian Research Council Centre for Excellence in Convergent Bio-Nano Science and Technology, University of Melbourne, Melbourne, VIC, Australia; Melbourne Sexual Health Centre and Department of Infectious Diseases, Alfred Hospital and Central Clinical School, Monash University, Melbourne, VIC, Australia

## Abstract

The capacity of antibodies to engage with innate and adaptive immune cells via the Fc region is important in preventing and controlling many infectious diseases, and is likely critical in SARS-CoV-2 infection. The evolution of such antibodies during convalescence from COVID-19 is largely unknown. We developed novel assays to measure Fc-dependent antibody functions against SARS-CoV-2 spike (S)-expressing cells in serial samples from a cohort of 53 subjects primarily with mild-moderate COVID-19, out to a maximum of 149 days post-infection. We found that S-specific antibodies capable of engaging dimeric FcγRIIa and FcγRIIIa decayed linearly over time. S-specific antibody-dependent cellular cytotoxicity (ADCC) and antibody-dependent phagocytosis (ADP) activity within plasma declined linearly as well, in line with the decay of S-specific IgG. Although there was significant decay in S-specific plasma ADCC and ADP activity, they remained readily detectable by all assays in 94% of our cohort at the last timepoint studied, in contrast with neutralisation activity which was only detectable in 70% of our cohort by the last timepoint. Our results suggest that Fc effector functions such as ADCC and ADP could contribute to the durability of SARS-CoV-2 immunity, particularly late in convalescence when neutralising antibodies have waned. Understanding the protective potential of antibody Fc effector functions is critical for defining the durability of immunity generated by infection or vaccination.

## Introduction

Most individuals who recover from COVID-19 develop binding and neutralising antibody responses against SARS-CoV-2 spike (S) protein (*1, 2*), with neutralising antibody responses generally targeted to the receptor-binding domain (RBD) of S (*3*). Passive transfer of neutralising monoclonal antibodies (mAbs) can protect animal models from subsequent SARS-CoV-2 challenge (*4-6*), suggesting neutralisation is likely to be a correlate of protection in humans (*7*). However, the duration of protection from re-infection in humans conferred by neutralising antibodies is not known. Several studies now show neutralising antibodies decline rapidly during early convalescence (*2, 8, 9*), with the magnitude of the antibody response positively correlating with disease severity (*10, 11*). Following mild COVID-19, many subjects mount modest neutralising antibody responses that decline to undetectable levels within 60 days, despite the maintenance of S- and RBD-specific IgG binding antibodies (*10*). Given that reported cases of SARS-CoV-2 re-infection have been rare to date, it is likely that immune responses beyond neutralisation contribute to SARS-CoV-2 protective immunity. Apart from direct virus neutralisation, antibodies can also mediate antiviral activity such as antibody-dependent cellular cytotoxicity (ADCC) and antibody-dependent phagocytosis (ADP) by engaging Fc gamma receptors (FcγR) on NK cells or phagocytes. Fc effector functions contribute to the prevention and control of other viral infections including HIV-1, influenza and Ebola (*12-14*). Butler et al. recently showed that SARS-CoV-2 RBD-specific antibodies within plasma could crosslink Fcγ receptors, and mediate ADP and antibody-dependent complement deposition (*15*). Importantly, two recent challenge studies demonstrated that certain RBD-specific mAbs rely on Fc effector functions to mediate protection against SARS-CoV-2 in mice (*16, 17*).

We previously reported that binding antibodies to SARS-CoV-2 S exhibit substantially longer half-lives than the neutralising antibody response (*8*), suggesting that Fc-mediated antibody function may extend the protective window beyond that inferred from neutralising activity alone. At present, analyses of Fc-mediated functions of SARS-CoV-2 antibodies within COVID-19 convalescent subjects have focussed upon cross-sectional analyses or short-term longitudinal studies up to 1-2 months post-symptom onset (*15, 18, 19*). We extend these findings and analyse Fc effector functions mediated by S-specific antibodies in a cohort of 53 convalescent individuals up to 149 days post-symptom onset. We developed novel functional assays using SARS-CoV-2 S-expressing cells to comprehensively analyse plasma ADCC and ADP activity against SARS-CoV-2 S. Our results show that plasma ADCC and ADP activity decay over the first 4 months post-infection, mirroring the decline in S-specific IgG titres. Importantly, however, S-specific antibodies capable of Fc-mediated antiviral activity remain readily detectable in almost all donors out to 4 months post-infection, even in donors whose neutralising antibody responses have waned to undetectable levels. Consequently, S-specific IgG could potentially mediate Fc-dependent effector functions that contribute to protection from SARS-CoV-2 infection even in the absence of plasma neutralising activity.

## Results

### Decay of dimeric FcγR-binding S and RBD-specific antibodies

We collected repeated (2-4) longitudinal samples from a cohort of 53 subjects after recovery from COVID-19 (Fig 1A, Table S1). The first sample was collected at a median of 41 days post-symptom onset (IQR 36-48) while the last sample was collected at a median of 123 days post-symptom onset (IQR 86-135). The engagement of dimeric recombinant soluble FcγRIIIa and FcγRIIa proteins by antibodies mimics the immunological synapse required for FcγR activation of innate immune cells, and is a surrogate measure of ADCC and ADP respectively (*20, 21*). To determine the dynamics of Fc-mediated function in plasma samples over time, we measured the capacity of dimeric FcγRIIIa and FcγRIIa receptors to engage antibodies specific for SARS-CoV-2 S antigens (trimeric S, S1 or S2 subunits or the RBD; Table S2). Using mixed-effects modelling, we assessed the fit of single-phase or two-phase decay in FcγR-binding between the timepoints analysed. We found that dimeric FcγRIIIa (V158)-binding antibodies against SARS-CoV-2 trimeric S and RBD both had single-phase decay kinetics with half-lives (*t*_½_) of 175 and 95 days respectively (Fig. 1B-C). Dimeric FcγRIIa (H131) binding-antibodies against SARS-CoV-2 trimeric S and RBD also decayed constantly with *t*_½_of 175 and 74 days respectively. Kinetics of decay for dimeric FcγR-binding antibodies against S and RBD for the lower affinity polymorphisms of FcγRIIIa (F158) and FcγRIIa (R131) were broadly similar to their higher affinity counterparts (Fig. S1A), with dimeric FcγR-binding antibodies against RBD decaying faster than for S. Consistent with our previous report that S1-specific IgG decays faster than S2-specific IgG(*8*), FcγR binding activity with antibodies against the S1 subunit decayed faster than that of S2 (FcγRIIIa, *t*_½_of 84 vs 227 days; FcγRIIa, *t*_½_of 65 vs 317 days; Fig. S1B).

**Fig 1.**
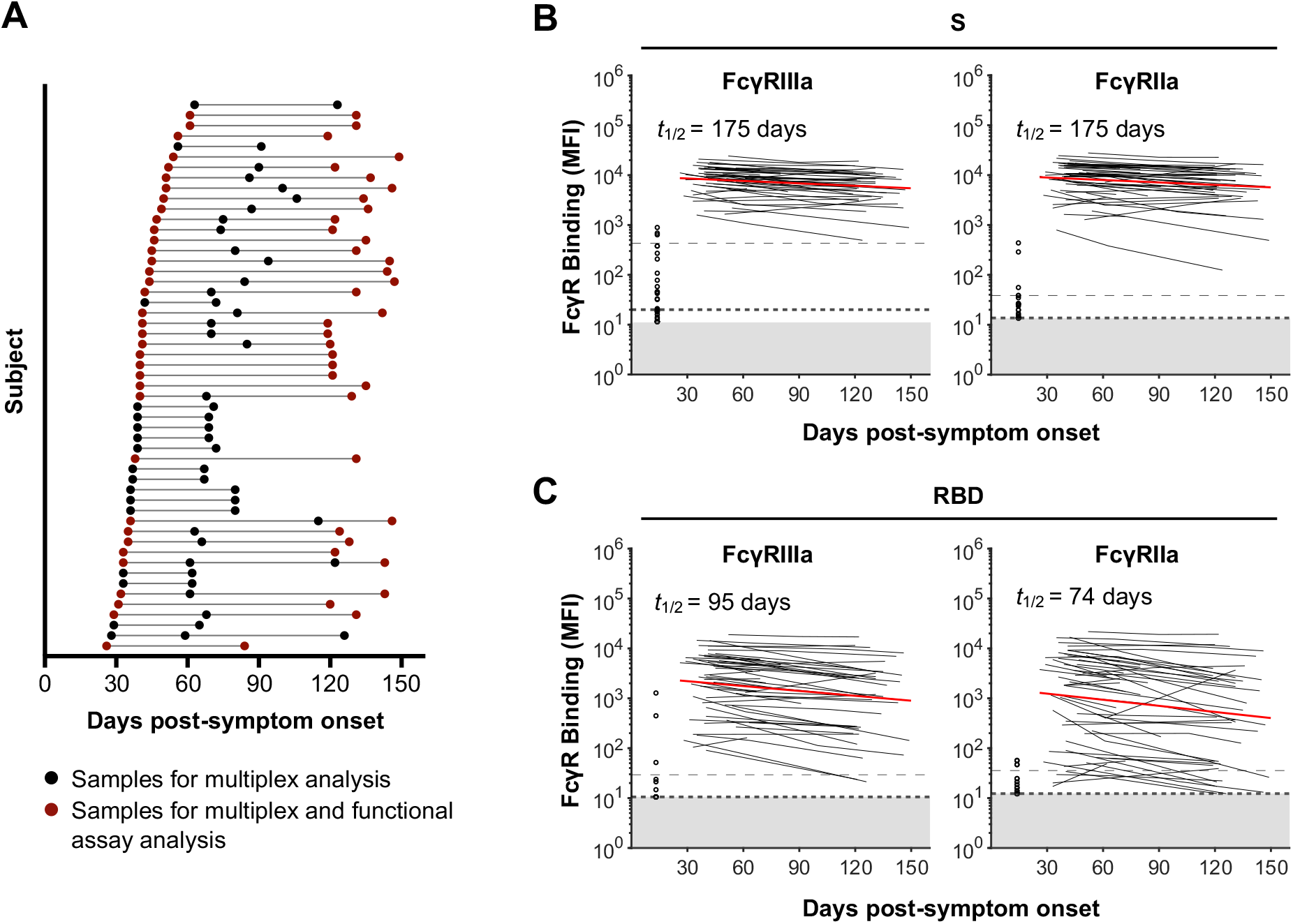
Dynamics of SARS-CoV-2 S and RBD-specific dimeric FcγR-binding antibodies in COVID-19 convalescent individuals. **(A)** Timeline of sample collection for each COVID-19 convalescent subject (n=53). Subjects with 2 samples at least 60 days apart were chosen for functional assay analysis (n=36). **(B-C)** Kinetics of SARS-CoV-2 S and RBD-specific dimeric FcγRIIIa (V158) and dimeric FcγRIIa (H131) binding antibodies over time. The best-fit decay slopes (red lines) and estimated half-lives (*t*_½_) are indicated for COVID-19 convalescent individuals. Uninfected controls (n=33) are shown in open circles, with the median and 90% percentile responses presented as thick and thin dashed lines respectively. The limit of detection is shown as the shaded area.

### Decay of S-specific ADCC

ADCC could play a role in eliminating cells infected with SARS-CoV-2. We generated Ramos- and A549-derived cell lines as model target cells that stably express membrane-localised S with either mOrange2 or luciferase reporters (Fig. S2A-B). The capacity of plasma IgG to recognise S was measured in 36 subjects in our cohort who had at least 60 days between the first and last visits (median of 89 days between first and last visits; Table S1) and 8 seronegative controls. Using a Ramos cell line expressing high levels of S (Ramos S-Orange) (Fig. S2C), we find IgG binding to cell-surface displayed S proteins decayed significantly between the first and last visits (*p*<0.0001; Fig. S2C) with a half-life of 97 days (Fig. S3). These results are consistent with the decay of S-specific IgG titres we observed previously (*8*) and the decay of dimeric FcγR-binding antibodies against S in Fig 1B.

As a surrogate measure of ADCC, we next used FcγRIIIa reporter cells to quantify the capacity of S-specific antibodies in plasma to engage cell surface FcγRIIIa and activate downstream NF-κB signalling (measured by induced nano-luciferase expression in the FcγRIIIa reporter cells) (Fig. 2A, Fig. S4A). FcγRIIIa activity decayed significantly over time (*p*<0.0001; Fig. 2C) with a half-life of 119 days (Fig. S3), and was correlated with S-specific IgG titres measured using stably transduced cells or by binding to dimeric FcγRIIIa (Fig. 2D). To confirm antibody recognition could mediate killing of S-expressing cells, we quantified the loss of cellular luciferase signal in Ramos S-luciferase target cells in the presence of convalescent plasma and primary human NK cells (Fig. 2B, Fig. S4B). S-specific ADCC decayed significantly over time (*p*<0.0001; Fig. 2E) with a half-life of 105 days (Fig. S3), and correlated with both cell-associated S-specific IgG and dimeric FcγRIIIa-binding antibodies against S (Fig. 2F).

**Fig 2.**
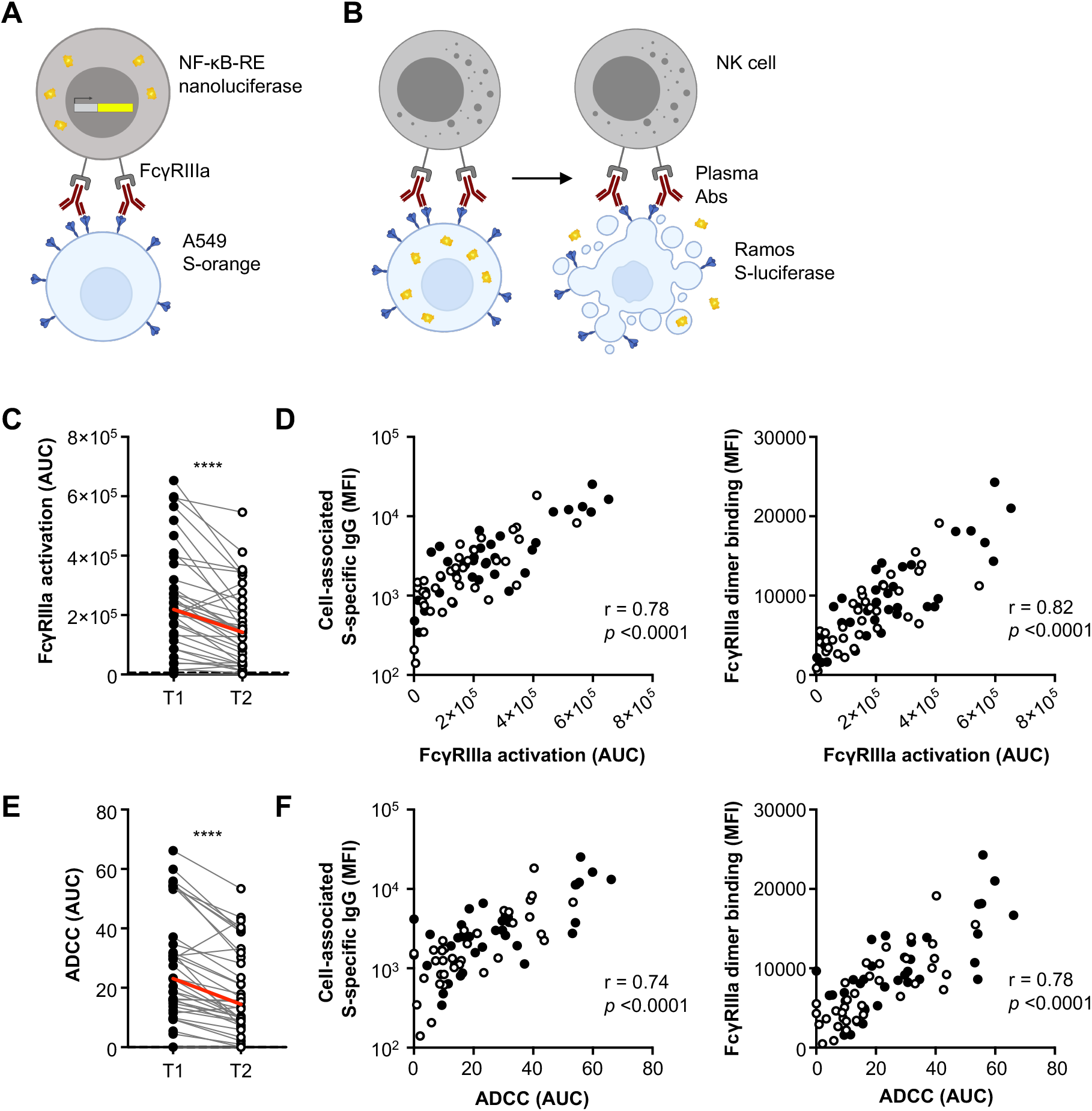
ADCC responses in COVID-19 convalescent individuals over time. **(A)** Schematic of the FcγRIIIa NF-κB activation assay. IIA1.6 cells expressing FcγRIIIa V158 and a NF-κB response element-driven nanoluciferase reporter were co-incubated with A549 S-orange target cells and plasma from COVID-19 convalescent individuals or uninfected controls. The engagement of FcγRIIIa by S-specific antibodies activates downstream NF-κB signalling and nano-luciferase expression. **(B)** Schematic of the luciferase-based ADCC assay. Purified NK cells from healthy donors were co-incubated with Ramos S-luciferase target cells and plasma. ADCC is measured as the loss of cellular luciferase. **(C)** S-specific FcγRIIIa-activating plasma antibodies in COVID-19 convalescent individuals in the first (T1; filled) and last (T2; open) timepoints available. **(D)** Correlation of S-specific FcγRIIIa-activating antibodies to cell-associated S-specific IgG and S-specific dimeric FcγRIIIa-binding antibodies. **(E)** S-specific ADCC mediated by plasma antibodies from COVID-19 convalescent individuals in the first (T1; filled) and last (T2; open) timepoints available. **(F)** Correlation of S-specific ADCC to cell-associated S-specific IgG and S-specific dimeric FcγRIIIa-binding antibodies. Red lines indicate the median responses of COVID-19 convalescent individuals (N=36) while dashed lines indicate median responses of uninfected controls (N=8). Statistical analyses were performed with the Wilcoxon signed-rank test (****, *p<*0.0001). Correlations were performed with the non-parametric Spearman test.

### Decay of S-specific ADP

As has been suggested for SARS-CoV, ADP could play a role in eliminating antibody-opsonised virions (*22*). We first used a well-established ADP assay (*23*) to measure antibody-mediated uptake of S-conjugated fluorescent beads into THP-1 monocytes (Fig. 3A; gating in Fig. S5A-B and optimisation in Fig. S6A-C). ADP of S-conjugated beads was detected in all 36 subjects at the first time point studied but decayed significantly over time (*p*<0.0001; Fig. 3C) with a half-life of 263 days (Fig. S3). ADP of S-conjugated beads correlated with cell-associated S-specific IgG and S-specific dimeric FcγRIIa-binding antibodies (Fig. 3D).

**Fig 3.**
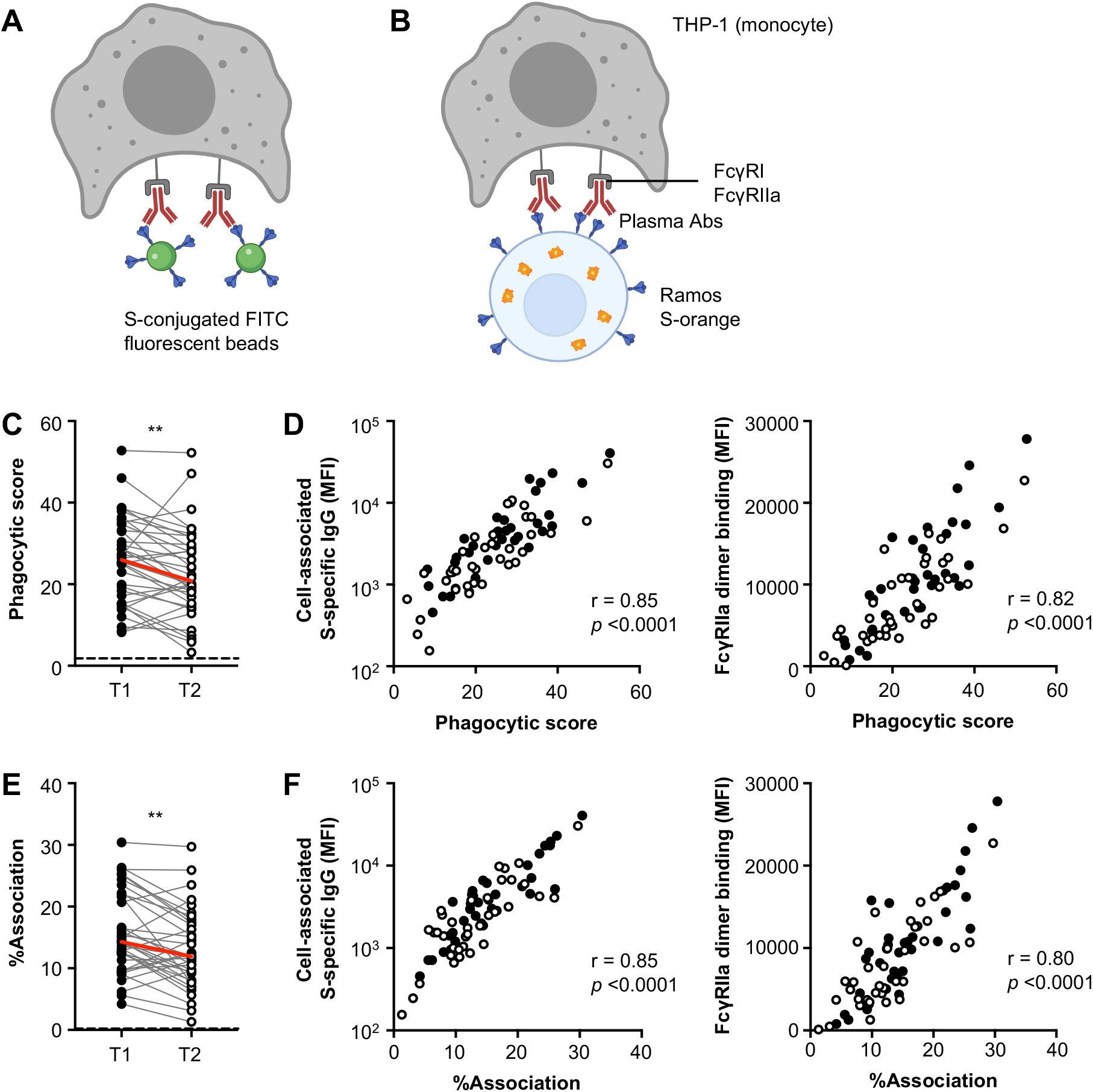
ADP responses in COVID-19 convalescent individuals over time. **(A)** Schematic of the bead-based ADP assay. THP-1 cells were incubated with S-conjugated fluorescent beads and plasma from COVID-19 convalescent individuals or uninfected controls. The uptake of fluorescent beads was measured by flow cytometry **(B)** Schematic of the THP-1 FcγR-dependent cell association assay. Ramos S-orange cells were pre-incubated with plasma prior to co-incubation with THP-1 cells. The association of THP-1 cells with Ramos S-orange cells was measured by flow cytometry. **(C)** ADP of S-conjugated beads mediated by plasma antibodies from COVID-19 convalescent individuals in the first (T1) and last (T2) timepoints available. **(D)** Correlation of ADP to cell-associated S-specific IgG and S-specific dimeric FcγRIIa-binding antibodies. **(E)** FcγR-dependent association of THP-1 cells with Ramos S-orange cells mediated by plasma antibodies from COVID-19 convalescent individuals in the first (T1) and last (T2) timepoints available. **(F)** Correlation of association of THP-1 cells with Ramos S-orange cells to cell-associated S-specific IgG and S-specific dimeric FcγRIIIa-binding antibodies. Red lines indicate the median responses of COVID-19 convalescent individuals (N=36) while dashed lines indicate median responses of uninfected controls (N=8). Statistical analyses were performed with the Wilcoxon signed-rank test (**, *p<*0.01). Correlations were performed with the non-parametric Spearman test.

In addition to uptake of antibody-opsonised virions, phagocytes could also potentially mediate clearance of infected cells expressing SARS-CoV-2 S on the cell surface. THP-1 cells have been shown to mediate both trogocytosis (sampling of plasma membrane fragments from target cells that can lead to cell death) and phagocytosis via antibody Fc-FcγR interactions with target cells (*24-26*). As such, we measured the FcγR-dependent association of THP-1 cells with Ramos S-orange cells following incubation with plasma from convalescent individuals or uninfected controls (Fig. 3B; gating in Fig. S5C and optimisation in Fig. S6D-F). Association of THP-1 cells with Ramos S-orange cells was detected in all subjects at the first time point but decayed significantly over time (*p*<0.0001; Fig. 3E) with a half-life of 351 days (Fig. S3), correlating with IgG binding to cell-associated S and S-specific dimeric FcγRIIa-binding antibodies (Fig. 3F).

### Cross-reactivity with HCoV S-specific antibodies

Cross-reactive antibodies between endemic human coronaviruses (HCoV) and SARS-CoV-2 have been widely reported (*27, 28*), suggesting past exposure to HCoVs may prime ADCC and ADP immunity against SARS-CoV-2. In addition, several studies have shown back-boosting of antibodies against endemic human coronaviruses (HCoV) following infection with SARS-CoV-2 (*29, 30*), likely due to the recall of pre-existing B cell responses against conserved regions of S. We thus determined whether IgG antibodies against S from four HCoV strains (OC43, HKU1, 229E and NL63) (Table S2) were boosted in COVID-19 convalescent subjects compared to uninfected healthy controls. We found that COVID-19 convalescent subjects had increased IgG antibodies against S from the betacoronaviruses OC43 and HKU1 (that are more closely related to SARS-CoV-2) at the first timepoint sampled compared to uninfected controls (Fig S7), while there was no difference in IgG levels against S from the alphacoronaviruses 229E and NL63. Correspondingly, the elevated IgG against OC43 and HKU1 S decayed over time while IgG against 229E and NL63 S remained stable (Fig 4A). We then measured whether dimeric FcγR-binding antibodies against HCoV S antigens in COVID-19 convalescent individuals declined over time. Dimeric FcγR-binding antibodies against OC43 and HKU1 S antigens were much higher in COVID-19 convalescent individuals than in healthy controls and decayed more rapidly over time compared to that against 229E and NL63 (Fig. 4A, Fig S8A-C). While there was an overall decay of dimeric FcγR-binding antibodies against OC43 S (FcγRIIIa *t*_½_= 224, FcγRIIa *t*_½_= 171 days), this was largely due to a decay in antibodies against the more conserved S2 subunit (FcγRIIIa *t*_½_= 229, FcγRIIa *t*_½_= 179 days) as FcγR-binding antibodies against the S1 subunit were not boosted and did not change substantially over time (Fig. 4B-C). This was also the case for HKU1, where dimeric FcγR-binding antibodies against S decayed over time but antibodies against the S1 subunit did not change (Fig S8A).

**Fig 4.**
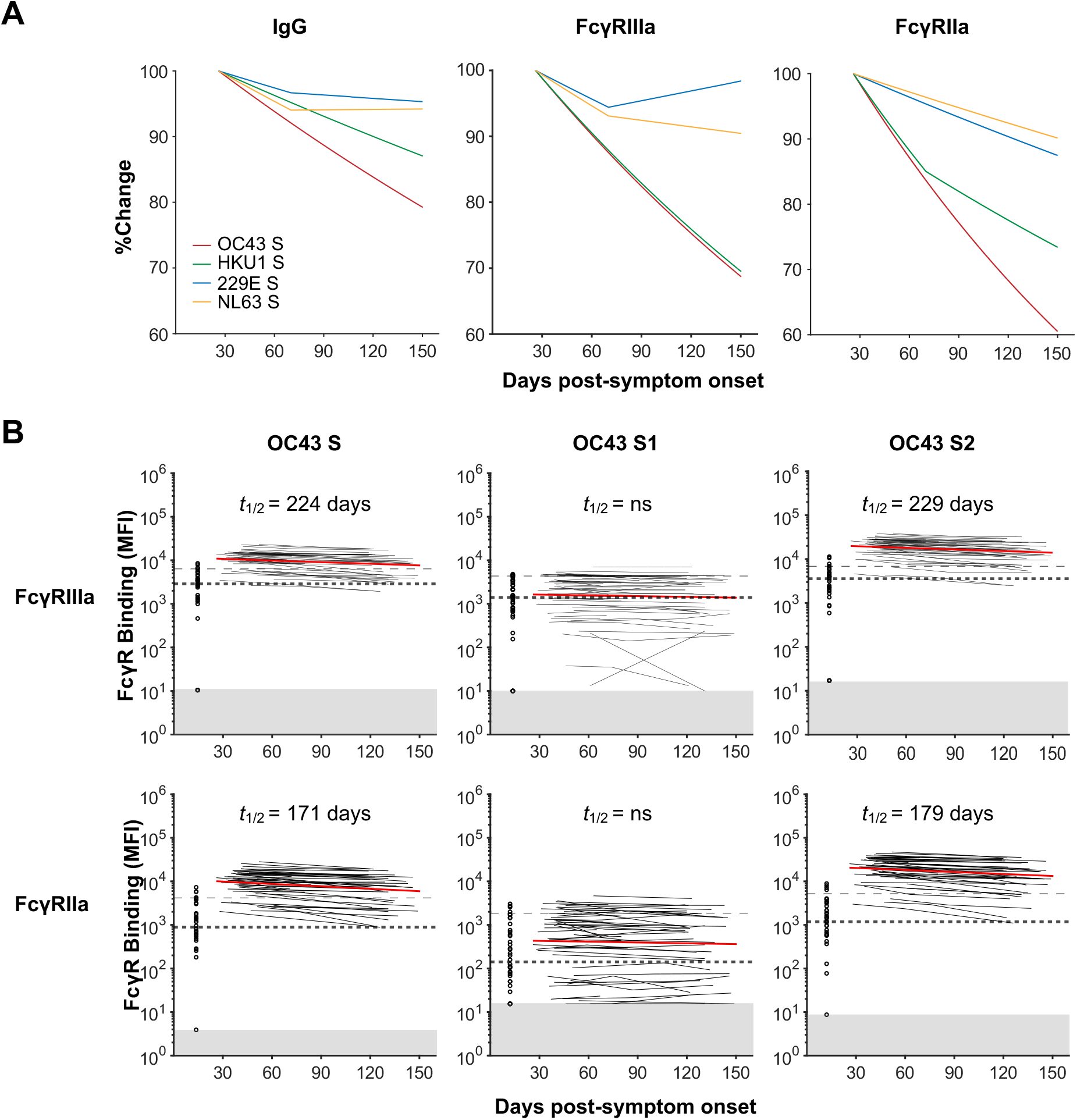
Dynamics of HCoV S-specific antibodies in COVID-19 convalescent individuals. **(A)** Best fit decay slopes of IgG and dimeric FcγR-binding antibodies against S from HCoV strains OC43, HKU1, 229E and NL63. The responses at timepoint 1 for each parameter are set to 100% and the %change over time is shown. **(B-C)** Kinetics of dimeric FcγRIIIa (V158) and FcγRIIa (H131) binding antibodies against HCoV-OC43 S antigens over time in COVID-19 convalescent individuals (n=53). The best-fit decay slopes (red lines) and estimated half-lives (*t*_½_) are indicated for COVID-19 convalescent individuals. Uninfected controls (n=33) are shown in open circles, with the median and 90% percentile responses presented as thick and thin dashed lines respectively. The limit of detection is shown as the shaded area.

### Decay kinetics of S-specific antibodies, neutralisation and Fc effector functions

To compare the decay kinetics of S-specific antibodies, neutralisation and Fc effector functions, we plotted the best fit decay slopes over time as a percentage of the response measured at timepoint 1 (Fig. 5A). The best-fit decay slopes of S-specific IgG and plasma neutralisation titres were obtained from a previous dataset that encompass the same subjects analysed for dimeric FcγR-binding antibodies and Fc effector functions (*8*) (Fig. S3). The general decline in plasma S-specific IgG titres and dimeric FcγR-binding activity was similarly reflected in reductions in Fc effector functions during convalescence from COVID-19. Importantly, Fc effector functions at the last timepoint sampled were still readily detectable above baseline activity observed in uninfected controls (97% for FcγRIIIa activation, 94% for ADCC, 100% for ADP and 100% for THP-1 association). This contrasted with plasma neutralisation activity, which was detectable above background for only 70% of subjects (Fig. 5B). The longer persistence of S-specific IgG and dimeric FcγR-binding antibodies against S has important implications as they may contribute to protection from SARS-CoV-2 infection following the decline of neutralising antibodies.

**Fig 5.**
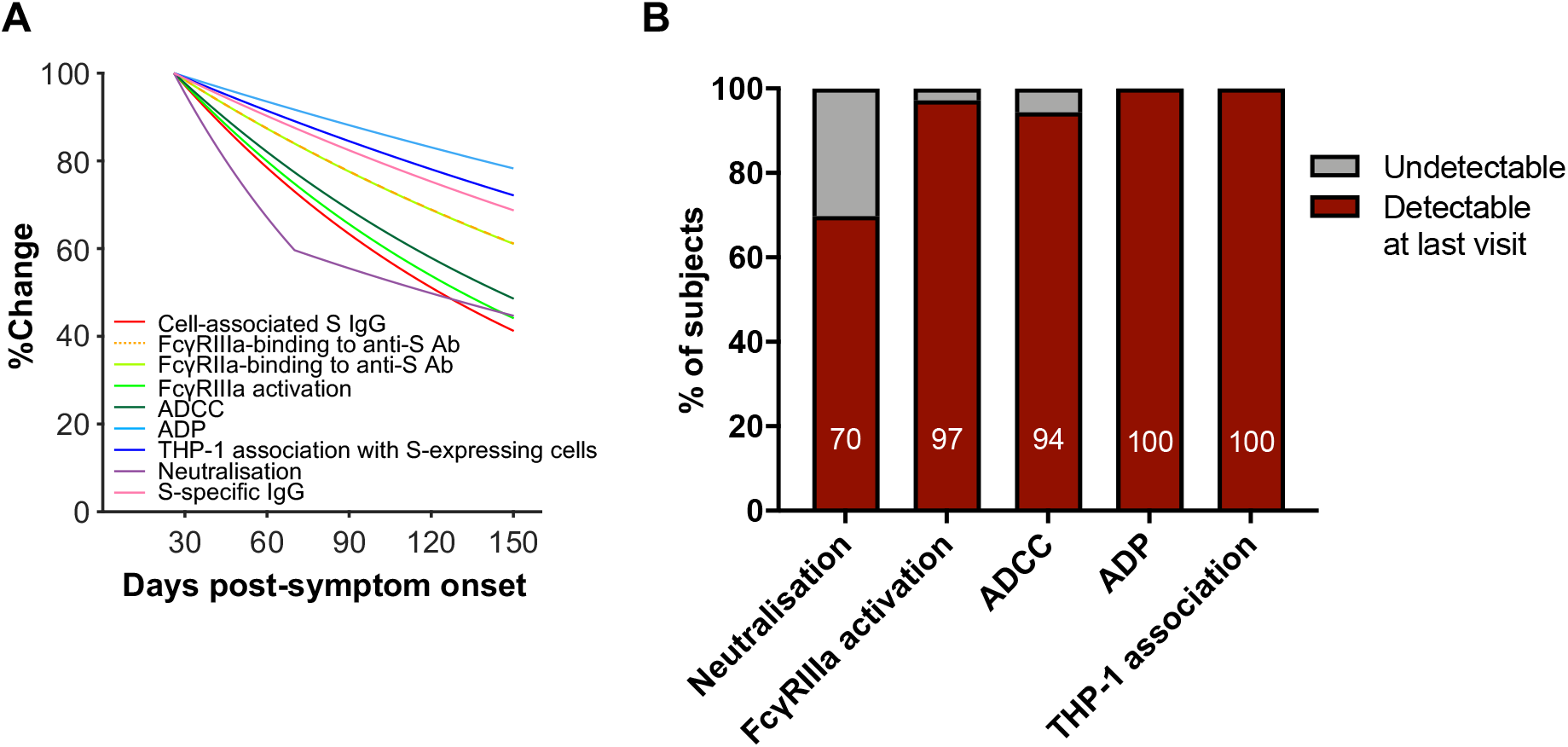
Decay kinetics of binding antibodies, neutralisation and Fc effector functions following SARS-CoV-2 infection. **(A)** Best fit decay slopes of various antibody parameters against SARS-CoV-2 S over time. The responses at timepoint 1 for each parameter are set to 100% and the %change over time is shown. **(B)** The percentage of subjects having detectable responses above (red) and below (grey) background levels at the last visit are shown. Background levels for each assay were the median responses of uninfected controls.

## Discussion

Using a multiplex bead array and novel assays measuring Fc effector functions against SARS-CoV-2 S, we find that FcγR-binding, ADCC and ADP activities of S-specific antibodies decay during convalescence from COVID-19. The decline of plasma ADCC and ADP activity correlated with the decay of S-specific IgG and FcγR-binding antibodies. Importantly, Fc effector functions were readily detectable above uninfected controls in 94% of subjects for all assays at the last timepoint sampled, in sharp contrast with neutralisation activity, which remained detectable above background for only 70% of subjects. While neutralising antibodies are likely to form a correlate of protection for SARS-CoV-2 (*7*), several studies find that neutralising antibodies in convalescent donors with mild COVID-19 wane rapidly (*2, 8, 9*). The rapid decline of plasma neutralisation activity in the early weeks following infection could potentially be explained by the rapid decline of plasma IgM and IgA titres against S and RBD (*19, 31*), which substantially contribute to neutralisation of SARS-CoV-2 (*32-34*). Given the relative scarcity of re-infection cases reported to date, it is likely that immune responses beyond neutralisation, including antibody Fc effector functions and T cell responses, contribute to long-term protection from SARS-CoV-2. Indeed, a recent study demonstrated that cellular immunity in convalescent macaques, mainly CD8^+^ T cells, contribute to protection against re-challenge after neutralising antibodies have waned (*22, 35*).

Our results demonstrate that FcγR-binding antibodies against betacoronaviruses OC43 and HKU1 are much higher in COVID-19 convalescent individuals compared to uninfected controls. This could either be due to the back-boosting of pre-existing HCoV antibodies that are cross-reactive with SARS-CoV-2 (*27, 28*), or the *de novo* generation of SARS-CoV-2 antibodies that are cross-reactive with conserved HCoV epitopes. Cross-reactive S antibodies were largely directed against the more conserved S2 subunit, in line with other reports (*27, 28*). A recent study found cross-reactive binding and neutralising antibodies against SARS-CoV-2 S2 in uninfected children and adolescents (*27*), suggesting prior infections with OC43 or HKU1 can elicit cross-reactive antibodies against the S2 subunit of SARS-CoV-2 S. These findings raise the interesting question of whether cross-reactive antibodies are recalled rapidly during early SARS-CoV-2 infection and can contribute to Fc effector functions against conserved epitopes within the S2 subunit. The presence of cross-reactive S2-specific antibodies capable of mediating Fc effector functions in early infection could potentially ameliorate disease symptoms and severity. Follow-up studies to dissect the influence of S1 or S2 antibody epitope localisation on FcγR engagement and the impact on Fc effector functions are also warranted.

Initial concerns for antibody-dependent enhancement (ADE) of COVID-19 were driven by the reported association of higher SARS-CoV-2 antibody titres with severe disease (*36*). However, this could simply be the result of prolonged antigen exposure due to higher viral loads. Importantly, Zohar et al. showed that in subjects with severe COVID-19, those who survived had higher levels of S-specific antibodies and Fc-mediated effector functions compared to those who died (*31*). Notably, numerous trials of convalescent plasma (CP) therapy for COVID-19 have been safely conducted (*37-39*), with no enhancement of disease reported to date (*40-42*). Since RBD-specific IgG1 antibodies in severe COVID-19 are more likely to have afucosylated Fc regions and trigger hyper-inflammatory responses from monocytes and macrophages (*43, 44*), there could be implications for ADE in people who are re-infected with SARS-CoV-2 after initial neutralising antibodies have waned but non-neutralising antibodies remain. Excessive Fc-mediated effector functions and immune complex formation in the absence of neutralisation could potentially trigger a hyper-inflammatory response and lead to ADE of disease, as observed for RSV and measles infections (*45, 46*). While ADE during re-infection remains only a theoretical risk, there have been two reported cases of re-infection where the second infection resulted in worse disease (*47, 48*). However, antibody levels after the first infection were not measured for one case (*47*) and only IgM was detectable after the first infection for the second case (*48*), arguing against Fc-mediated effector functions as the cause of increased pathogenicity.

Overall, we find that FcγR-binding, ADCC and ADP antibody functions decay following recovery from COVID-19 at a slower rate than serum neutralisation activity, suggesting non-neutralising antibody responses elicited by infection or vaccination may contribute to durable protection against SARS-CoV-2.

## Materials and methods

### Cohort recruitment and sample collection

People who recovered from COVID-19 and healthy controls were recruited to provide serial whole blood samples. Convalescent donors either had a PCR+ test during early infection or clear exposure to SARS-CoV-2, and were confirmed to have SARS-CoV-2 S- and RBD-specific antibodies via ELISA as previously reported (*1*). Contemporaneous uninfected controls who did not experience any COVID-19 symptoms were also recruited and confirmed to be seronegative via ELISA. For all subjects, whole blood was collected with sodium heparin or lithium heparin anticoagulant. The plasma fraction was then collected and stored at -80°C. A subset of 36 donors with at least 60 days between the first and last visits were chosen to proceed with the more labour-intensive functional ADCC and ADP assays. Plasma was heat-inactivated at 56°C for 30 minutes prior to use in functional assays. Characteristics of the COVID-19 convalescent and uninfected donors are described in Table S1. The study protocols were approved by the University of Melbourne Human Research Ethics Committee (#2056689). All subjects provided written informed consent in accordance with the Declaration of Helsinki.

### Luminex bead-based multiplex assay

As previously described (*49*), a custom multiplex bead array was designed and coupled with SARS-CoV-2 S trimer, S1 subunit (Sino Biological), S2 subunit (ACRO Biosystems) and RBD (BEI Resources), as well as HCoV (OC43, HKU1, 229E, NL63) S (Sino Biological) (Table S2). Tetanus toxoid (Sigma-Aldrich), influenza hemagglutinin (H1Cal2009; Sino Biological) and SIV gp120 (Sino Biological) were also included in the assay as positive and negative controls respectively. Antigens were covalently coupled to magnetic carboxylated beads (Bio Rad) using a two-step carbodiimide reaction and blocked with 0.1% BSA, before being resuspended and stored in PBS 0.05% sodium azide till use.

Using the coupled beads, a custom CoV multiplex assay was formed to investigate the dimeric recombinant soluble FcγR-binding capacity of pathogen-specific antibodies present in COVID-19 convalescent plasma samples and uninfected controls (*49*). Briefly, 20µl of working bead mixture (1000 beads per bead region) and 20µl of diluted plasma (final dilution 1:200) were added per well and incubated overnight at 4°C on a shaker. Different detectors were used to assess pathogen-specific antibodies. Single-step detection was done using phycoerythrin (PE)-conjugated mouse anti-human pan-IgG (Southern Biotech; 1.3µg/ml, 25µl/well). For the detection of FcγR-binding, recombinant soluble FcγR dimers (higher affinity polymorphisms FcγRIIIa-V158 and FcγRIIa-H131, lower affinity polymorphisms FcγRIIIa-F158 and FcγRIIa-R131; 1.3µg/ml, 25µl/well) were first added to the beads, washed, followed by the addition of streptavidin R-PE (Thermo Fisher Scientific). Assays were read on the Flexmap 3D and performed in duplicates.

### Cell lines

As target cells for the functional antibody assays, Ramos and A549 cells stably expressing full-length SARS-CoV-2 S and the reporter proteins mOrange2 or luciferase were generated by lentiviral transduction (Fig. S2A). To stain for S-expression, transduced cells were incubated with convalescent plasma (1:100 dilution) prior to staining with a secondary mouse anti-human IgG-APC antibody (1:200 dilution; clone HP6017, BioLegend). S-luciferase cells were bulk sorted on high S expression while S-orange cells were bulk sorted on high S- and mOrange2-expression. Following a week of outgrowth, the bulk sorted cells were single-cell sorted to obtain clonal populations of S-orange and S-luciferase cells (Fig. S2B). The Ramos cell lines were grown in complete RPMI medium (10% fetal calf serum (FCS) with 1% penicillin strepytomycin glutamine (PSG)) while the A549 cell lines were grown in complete DMEM medium (10% FCS with 1% PSG).

FcγRIIIa-NF-κB-RE nanoluciferase reporter cells were used as effector cells for the FcγRIIIa activation assay. IIA1.6 cells expressing the Fc receptor gamma subunit (FcR-γ) were maintained in RPMI containing 10% FCS, 2.5 mM L-glutamine, 55 µM 2-mercaptoethanol, 100 units penicillin and 100 units streptomycin (Sigma Aldrich). These were further transduced as described previously (*50*) using a FcγRIIIa V158 cDNA in pMX-neo and the packaging line Phoenix. IIA1.6/FcR-γ/FcγRIIIa V158 cells were transfected with a NF-κB response element driven nanoluciferase (NanoLuc) reporter construct (pNL3.2.NF-κB-RE[NlucP/NF-κB-RE/Hygro] (Promega) by nucleofection (Amaxa Kit T, Lonza) and selected in the presence of 200 µg/ml hygromycin. Reporter cells were maintained in media containing 400 µg/ml neomycin and 50 µg/ml hygromycin (ThermoFisher).

THP-1 monocytes (ATCC) were cultured in complete RPMI medium and maintained below a cell density of 0.3×10^6^/ml. Flow cytometry was used to confirm stable expression of FcγRIIa (CD32), FcγRI (CD64) and FcαR (CD89) on THP-1 monocytes prior to use in assays.

### FcγRIIIa activation assay

A549 S-orange cells were plated (2×10^5^/ml, 100 µl/well) in 96-well white flat-bottom plates (Corning). The next day, COVID-19 convalescent and uninfected plasma were serially diluted and 50 µl aliquots transferred to the aspirated A549 S-orange cells and incubated at 37°C, 60 min, 5% CO_2_. Unbound antibody was removed by aspirating the wells and refilling with RPMI (200 µl) four times. FcγRIIIa-NF-κB-RE nanoluciferase reporter cells (4 x 10^5^/ml, 50 µl/well) were added to the aspirated wells containing the opsonised A549 S-orange cells. After incubation (37^0^C, 4h, 5%CO_2_) cells were lysed by adding 50 µl/well of 10 mM Tris-pH 7.4, containing 5 mM EDTA, 0.5 mM DTT, 0.2% Igepal CA-630 (Sigma Aldrich), and Nano-Glo luciferase assay substrate (1:1000). Induction of nanoluciferase was measured using a 1s read on a Clariostar Optima plate reader (BMG Labtech) with background luminescence from control wells without agonist subtracted from test values.

### Luciferase-based ADCC assay

A luciferase-based ADCC assay was performed to examine ADCC against S-expressing cells. NK cells from healthy donors were first enriched from freshly isolated PBMCs using the EasySep Human NK Cell Enrichment Kit (Stemcell Technologies). In a 96-well V-bottom cell culture plate, purified NK cells (20,000/well) were mixed with Ramos S-luciferase cells (5,000/well) in the presence or absence of plasma from convalescent or uninfected donors at 1:100, 1:400 and 1:1600 dilutions. Each condition was tested in duplicate and “no plasma” and “target cell only” controls were included. Cells were centrifuged at 250g for 4 min prior to a 4-hour incubation at 37°C with 5% CO_2_. Cells were then washed with PBS and lysed with 25µl of passive lysis buffer (Promega). Cell lysates (20µl) were transferred to a white flat-bottom plate and developed with 30µl of britelite plus luciferase reagent (Perkin Elmer). Luminescence was read using a FLUOstar Omega microplate reader (BMG Labtech). The relative light units (RLU) measured were used to calculate %ADCC with the following formula: (“no plasma control” – “plasma sample”) ÷ “target cell only control” × 100. For each plasma sample, %ADCC was plotted against log_10_(plasma dilution^-1^) and the area under curve (AUC) was calculated using Graphpad Prism.

### Bead-based THP-1 ADP assay

To examine ADP mediated by COVID-19 convalescent plasma, a previously published bead-based ADP assay was adapted for use in the context of SARS-CoV-2 (*23*). SARS-CoV-2 S trimer was biotinylated using EZ-Link Sulfo-NHS-LC biotinylation kit (Thermo Scientific) with 20mmol excess according to manufacturer’s instructions and buffer exchanged using 30kDa Amicon centrifugal filters (EMD millipore) to remove free biotin. The binding sites of 1μm fluorescent NeutrAvidin Fluospheres beads (Invitrogen) were coated with biotinylated S at a 1:3 ratio overnight at 4°C. S-conjugated beads were washed four times with 2% BSA/PBS to remove excess antigen and incubated with plasma (1:100 dilution) for 2 hours at 37°C in a 96-well U-bottom plate (see Fig. S6 for optimisation). THP-1 monocytes (10,000/well) were then added to opsonized beads and incubated for 16 hours under cell culture conditions. Cells were fixed with 2% formaldehyde and acquired on a BD LSR Fortessa with a HTS. The data was analyzed using FlowJo 10.7.1 (see Fig. S5 for gating strategy) and a phagocytosis score was calculated as previously described (*51*) using the formula: (%bead-positive cells × mean fluorescent intensity)/10^3^. To account for non-specific uptake of S-conjugated beads, the phagocytosis scores for each plasma sample were subtracted with that of the “no plasma” control.

### Cell-based THP-1 association assay

To assess the capacity of THP-1 monocytes to associate with S-expressing target cells via Ab-FcγR interactions, an assay using THP-1 cells as effectors and Ramos S-orange cells as targets was performed. THP-1 monocytes were first stained with CellTrace^™^ Violet (CTV) (Life Technologies) as per manufacturer’s instructions. In a 96-well V-bottom cell culture plate, Ramos S-orange cells (10,000/well) were incubated with plasma from convalescent or uninfected donors (1:2700 dilution) for 30 minutes (see Fig. S6 for optimisation). Opsonised Ramos S-orange cells were then washed prior to co-culture with CTV-stained THP-1 monocytes (10,000/well) for 1 hour at 37°C with 5% CO_2_. After the incubation, cells were washed with PBS, fixed with 2% formaldehyde and acquired using the BD LSR Fortessa with a high-throughput sampler attachment (HTS). The data was analyzed using FlowJo 10.7.1 (see Fig. S5 for gating strategy). The percentage of Ramos S-orange cells associated with THP-1 monocytes (% association) was measured for each plasma sample and background-subtracted with the “no plasma” control.

### Decay rate estimation

The decay rate was estimated by fitting a linear mixed effect model for each response variable (y_ij_ for subject i at timepoint j) as a function of days post-symptom onset and assay replicate (as a binary categorical variable). The model can be written as below: *y*_*ij*_ *= β*_0_ + *b*_0*i*_ + *β*_1_*R*_*ij*_ + *β*_2_*t*_*ij*_+ *b*_2*i*_ *t*_*ij*_ – for a model with a single slope; and *y*_*ij*_ *= β*_0_ + *b*_0*i*_ + *β*_1_*R*_*ij*_ + *β*_2_*t*_*ij*_ + *b*_2*i*_ *t*_*ij*_ + *β*_3_ *s*_*ij*_ + *b*_3*i*_ *s*_*ij*_ – for a model with two different slopes, in which:

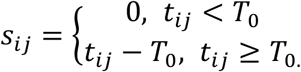

The parameter *β*_0_ is a constant (intercept), and *b*_0*i*_ is a subject-specific adjustment to the overall intercept. The slope parameter *β*_2_ is a fixed effect to capture the decay slope before *T*_0_ (as a fixed parameter, 70 days); which also has a subject-specific random effect *b*_2*i*_. To fit a model with two different decay rates, an extra parameter *β*_3_ (with a subject-specific random effect *b*_3*i*_) was added to represent the difference between the two slopes. Assay variability between replicates (only for HCoV response variables) was modelled as a single fixed effect *β*_1_, in which we coded the replicate as a binary categorical variable *R*_*ij*_. The random effect was assumed to be normally distributed with zero mean and variance *δ*.

We fitted the model to log-transformed data of various response variables, and we censored the data from below if it was less than the threshold for detection. The response variables had background levels subtracted by taking the mean of all the background values, and the threshold for detection was set at two standard deviations of the background responses. The model was fitted by using *lmec* library in *R*, using the ML algorithm to fit for the fixed effects. We also tested if the response variables can be fitted better by using a single or two different decay slopes (likelihood ratio test – based on the likelihood value and the difference in the number of parameters). These analyses were carried out in *R: A language and environment for statistical computing* version 4.0.2.

### Statistics

Statistical analyses were performed with Graphpad Prism 8. Correlations between functional ADCC and ADP responses with cell-associated S-specific IgG and FcγR-binding S-specific antibodies were assessed using the non-parametric Spearman test. Comparisons of functional ADCC and ADP responses between first and last visits were performed using the Wilcoxon signed-rank test. Comparisons between uninfected individuals and COVID-19 convalescent individuals were performed using the Mann-Whitney test.

## Data Availability

All data is available from the authors upon reasonable request

## Acknowledgments

We thank the cohort participants for generously providing samples. We thank Francesca Mordant and Kanta Subbarao (University of Melbourne) for performing the SARS-CoV-2 neutralisation assays. We acknowledge the Melbourne Cytometry Platform for provision of flow cytometry services. The following reagent was produced under HHSN272201400008C and obtained through BEI Resources, NIAID, NIH: Spike Glycoprotein Receptor Binding Domain (RBD) from SARS-Related Coronavirus 2, Wuhan-Hu-1 with C-Terminal Histidine Tag, Recombinant from HEK293F Cells, NR-52366. This study was supported by the Victorian Government, an Australian government Medical Research Future Fund award GNT2002073 (SJK, MPD, and AKW), the ARC Centre of Excellence in Convergent Bio-Nano Science and Technology (SJK), an NHMRC program grant APP1149990 (SJK and MPD), NHMRC project grants GNT1162760 (AKW), GNT1145303 (PMH and BDW), an NHMRC-EU collaborative award APP1115828 (SJK and MPD), the European Union Horizon 2020 Research and Innovation Programme under grant agreement 681137 (SJK), and Emergent Ventures Fast Grants (AWC). JAJ, AWC and SJK are supported by NHMRC fellowships. AKW, DC and MPD are supported by NHMRC Investigator grants. Figures were created using BioRender.

