## Supplementary Figures and Tables for "Decay of Fc-dependent antibody functions after mild to moderate COVID-19"

### Supplementary materials

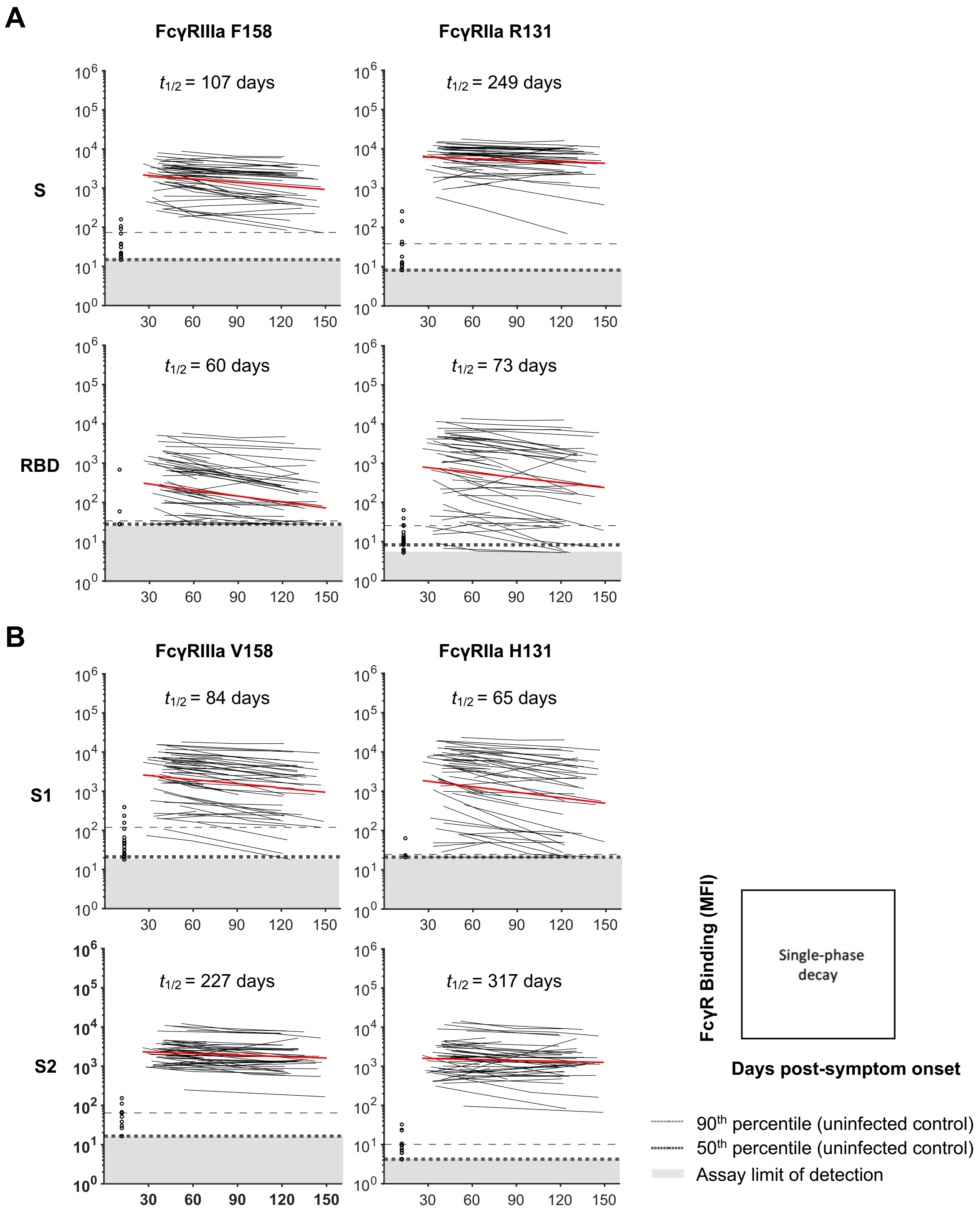


**Fig. S1. Dynamics of dimeric FcγR-binding antibodies against SARS-CoV-2 S antigens (S, S1, S2 and RBD) in COVID-19 convalescent individuals. (A)** Kinetics of SARS-CoV-2 S- and RBD-specific dimeric FcγRIIIa (F158) and dimeric FcγRIIa (R131)-binding antibodies over time. **(B)** Kinetics of SARS-CoV-2 S1 and S2 subunitspecific dimeric FcγRIIIa (V158) and FcγRIIa (H131) binding antibodies over time. The best-fit decay slopes (red lines) and estimated half-lives ($t_{1/2}$) are indicated for COVID-19 convalescent individuals. Uninfected controls (n=33) are shown in open circles, with the median and 90% percentile responses presented as thick and thin dashed lines respectively. The limit of detection is shown as the shaded area.

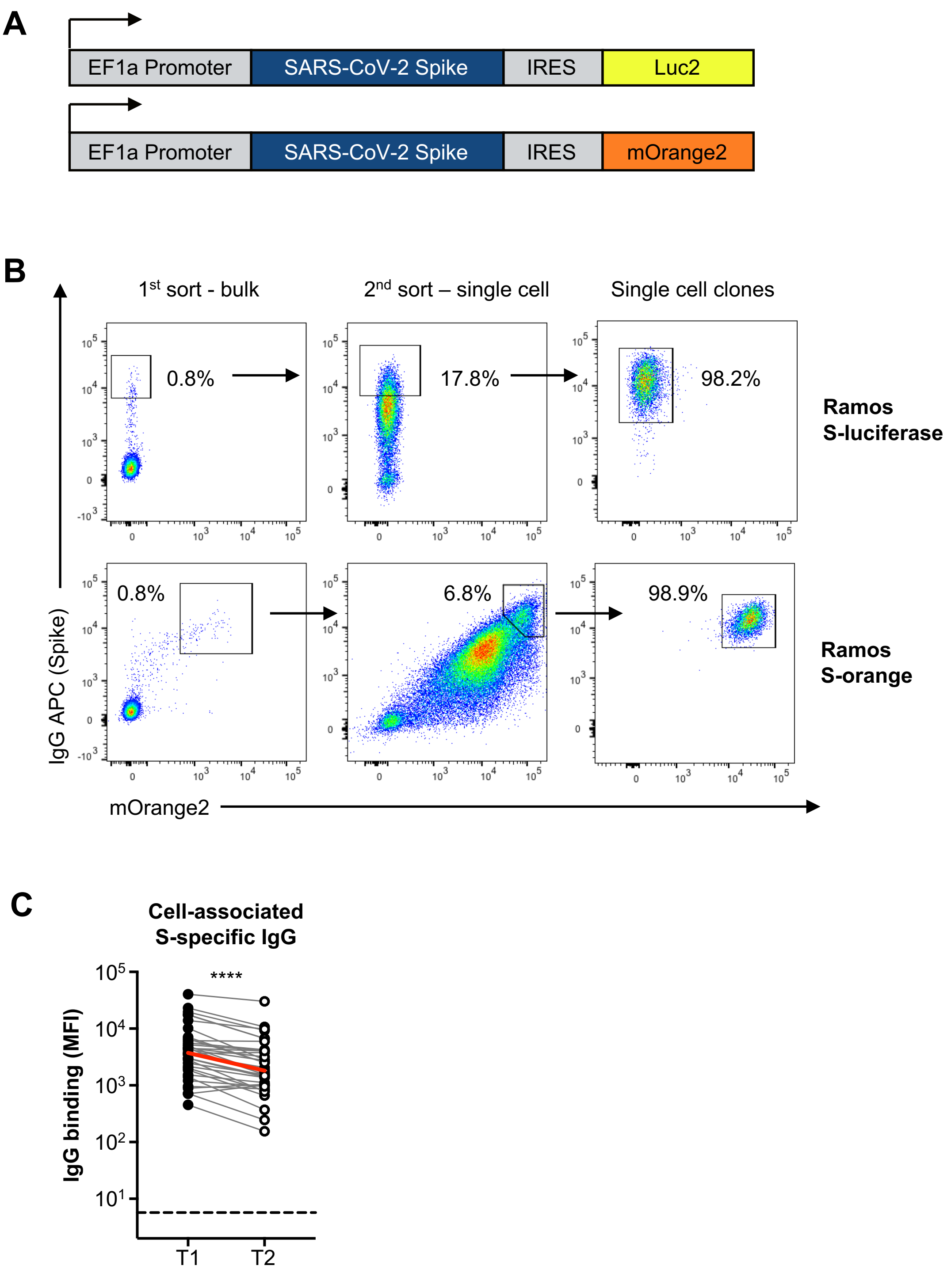


**Fig. S2. Generation of stable S-expressing reporter cell lines. (A)** Schematic of the constructs used to generate Ramos and A549 cell lines stably expressing SARS-CoV-2 S and the reporter proteins luciferase or mOrange2. **(B)** S-luciferase transduced cells were sorted on high S-expression while S-orange transduced cells were sorted on high S- and mOrange2-expression. Transduced cells were first bulk sorted, outgrown and then single cell sorted to obtain single cell clonal populations. **(C)** IgG binding to S expressed on Ramos S-orange cells within plasma from COVID-19 convalescent individuals in the first (T1) and last (T2) timepoints available. Red lines indicate the median responses of COVID-19 convalescent individuals (N=36) while dashed lines indicate median responses of uninfected controls (N=8). Statistical analyses between matched samples were performed with a Wilcoxon signed-rank test (****, $p$<0.0001)

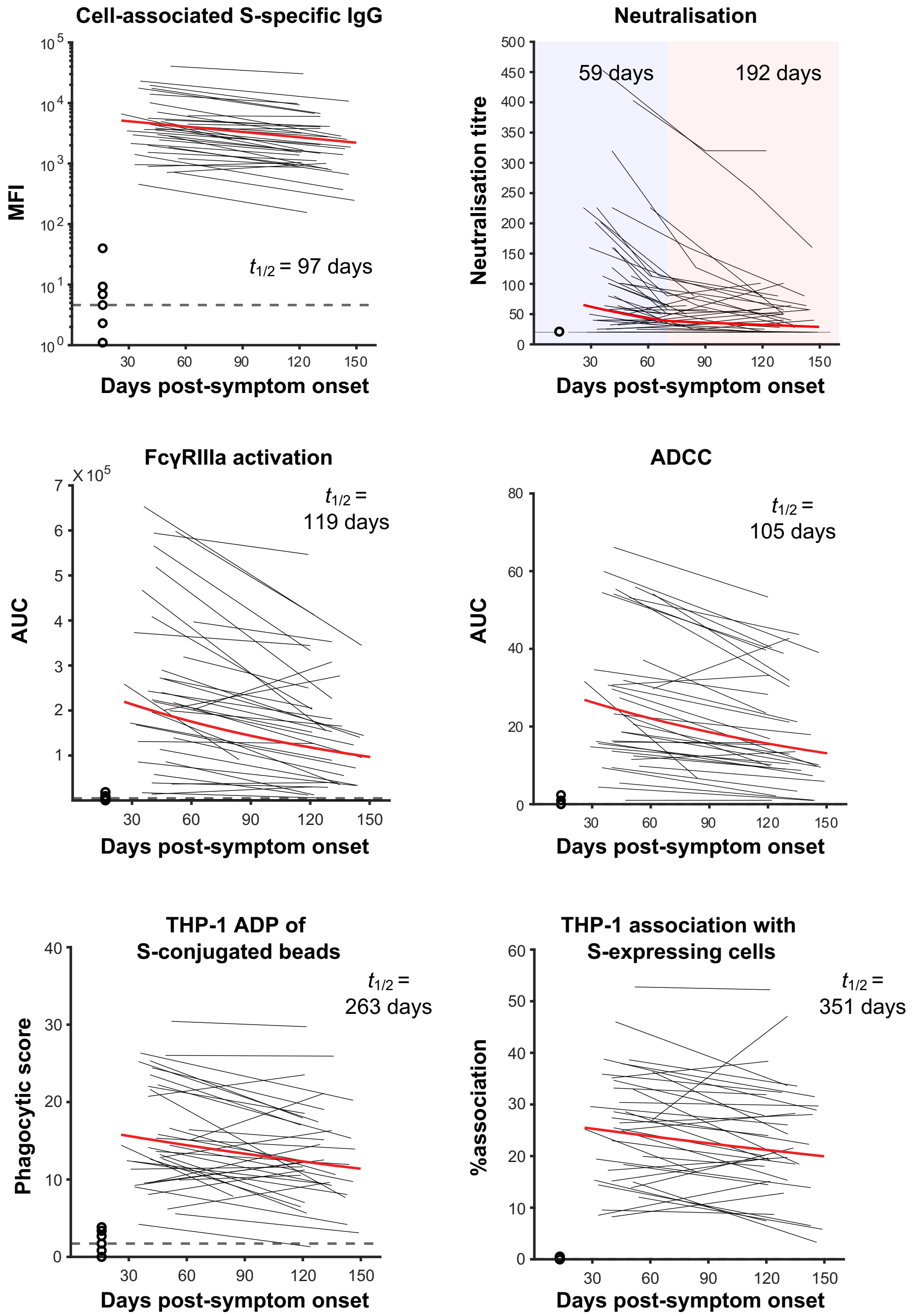


**Fig. S3. Fitting of SARS-CoV-2 cell-associated S-specific IgG, neutralisation and Fc effector functions over time.** The best-fit decay slopes (red lines) and estimated half-lives ($t_{1/2}$) are indicated for COVID-19 convalescent individuals. Uninfected controls are shown in open circles, with the median response presented as a dashed line.

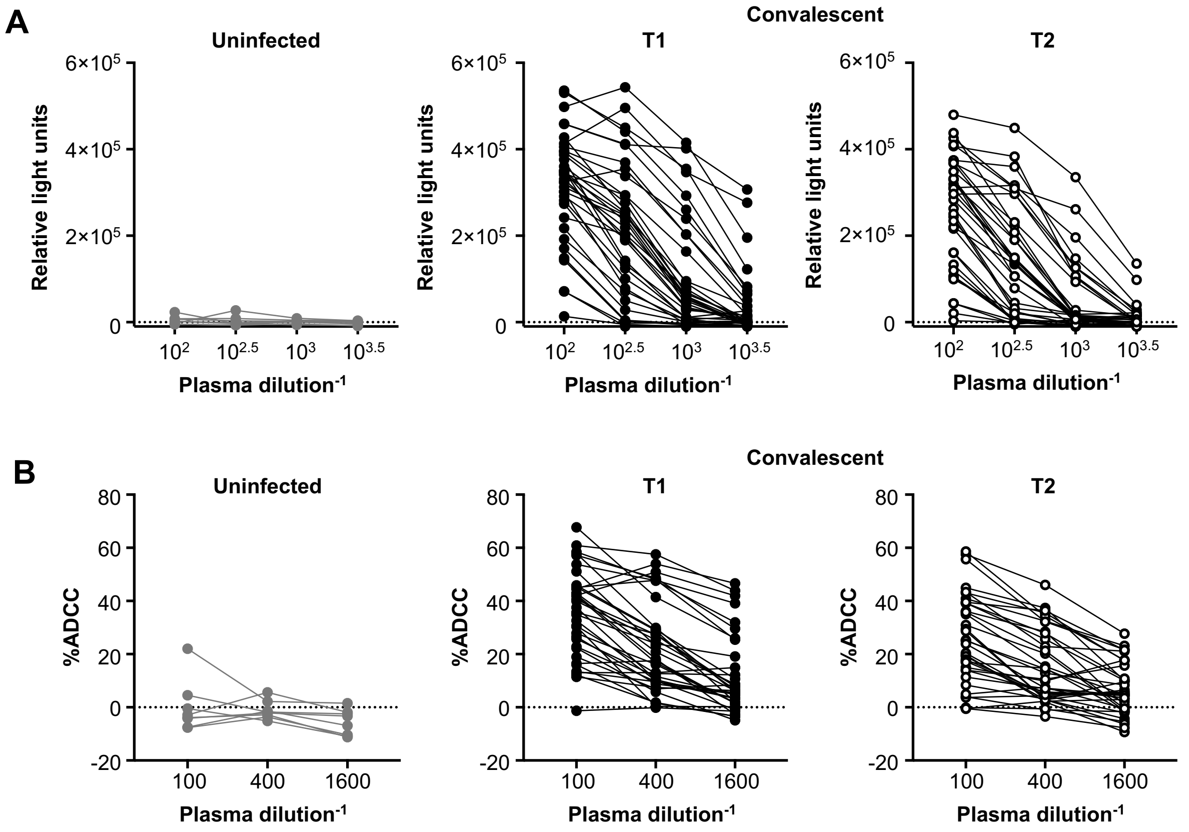


**Fig. S4. Dilution curves for the (A) FcγRIIIa NF-κB activation and (B) luciferase-based ADCC assays.** Plasma from uninfected controls (n=8) are shown in grey and plasma from COVID-19 convalescent donors (n=36) are shown in black, with the first available timepoint shown in filled circles (T1) and last available timepoint shown in open circles (T2). The area under curve (AUC) for each plasma donor was calculated and plotted in Fig. 2C and 2E.

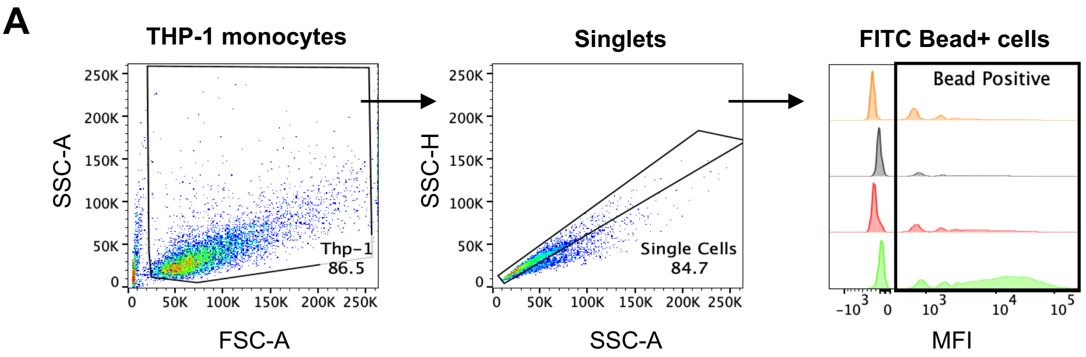


**B**

| | Sample ID | Bead Positive Cells | | Phagocytosis score |
|---|---|---|---|---|
| | | MFI | % | |
| (orange) | Uncoated beads | 1894 | 47.9 | 9.07 |
| (grey) | S-trimer beads – no plasma | 1822 | 37.1 | 6.76 |
| (red) | S-trimer beads – uninfected plasma | 1907 | 39.4 | 7.51 |
| (green) | S-trimer beads – convalescent plasma | 9374 | 77.4 | 72.55 |

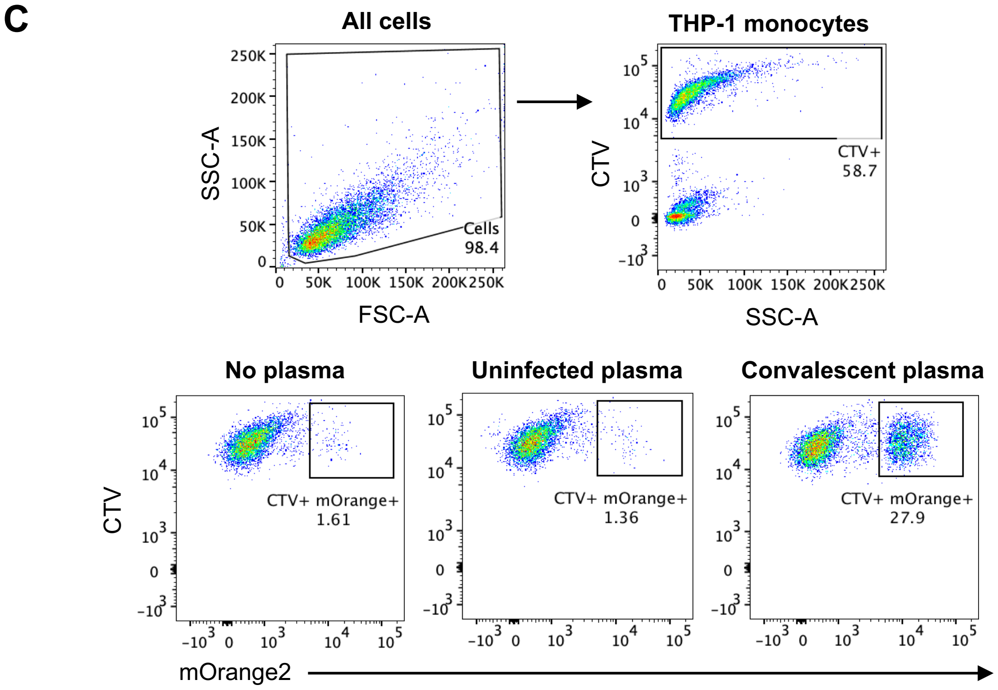


**Fig. S5. Gating strategy for THP-1 bead-based ADP and cell-association assays.** **(A)** After selection of THP-1 monocytes to remove debris (SSC-A vs FSC-A), a single cell gate was applied to exclude doublets (SSC-H vs SSC-A). Using histograms, THP-

1 monocytes were gated for uptake of S-conjugated or unconjugated FITC fluorescent beads. **(B)** The geometric mean fluorescent intensity (MFI) and the percentage of FITC bead+ cells (%) were multiplied and divided by $10^3$ to give an arbitrary phagocytosis score that can be used to compare conditions. **(C)** After selection of all cells to remove debris (SSC-A vs FSC-A), CellTrace Violet-stained THP-1 monocytes were gated (CTV vs SSC-A) and assessed for association with Ramos S-orange cells based on mOrange2 fluorescence. The bottom panels are representative plots of THP-1 cells associating with Ramos S-orange cells in the presence of no plasma, plasma from an uninfected control and plasma from a COVID-19 convalescent donor.

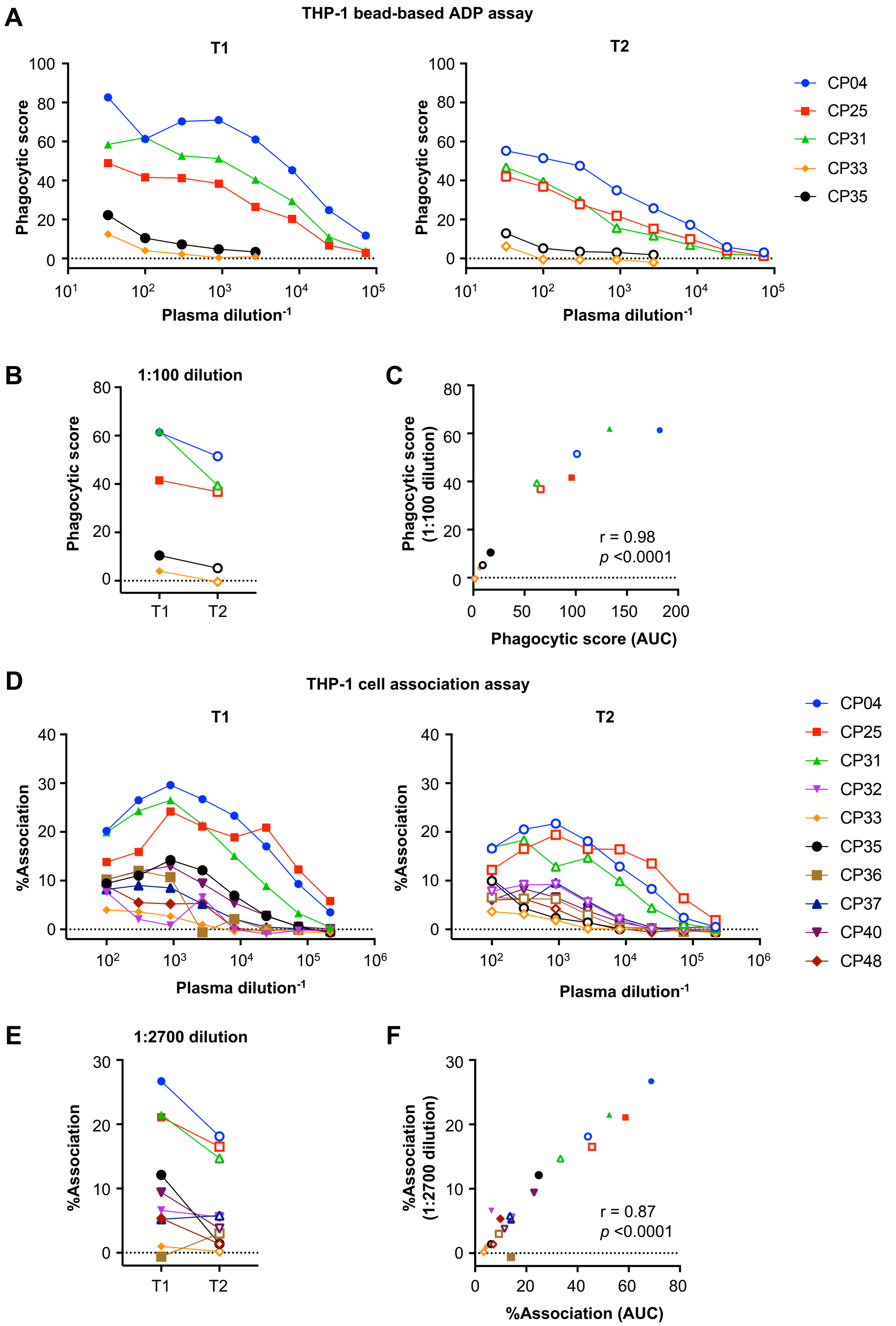
A
THP-1 bead-based ADP assay
T1
T2
Phagocytic score
Plasma dilution$^{-1}$
CP04
CP25
CP31
CP33
CP35
B
1:100 dilution
Phagocytic score
T1
T2
C
Phagocytic score (1:100 dilution)
r = 0.98
$p$ <0.0001
Phagocytic score (AUC)
D
THP-1 cell association assay
T1
T2
%Association
Plasma dilution$^{-1}$
CP04
CP25
CP31
CP32
CP33
CP35
CP36
CP37
CP40
CP48
E
1:2700 dilution
%Association
T1
T2
F
%Association (1:2700 dilution)
r = 0.87
$p$ <0.0001
%Association (AUC)

74

**Fig. S6. Optimisation of THP-1 bead-based ADP and cell-association assays. (A)** Using the THP-1 bead-based ADP assay, a 3-fold titration series was performed on a subset of 5 COVID-19 convalescent donors. **(B)** A 1:100 plasma dilution was chosen as the optimal dilution since a decay in the phagocytic score could be observed from timepoint 1 to timepoint 2 and **(C)** phagocytic score at 1:100 correlated significantly with AUC of the titration curves. **(D)** Using the THP-1 and Ramos S-orange cell association assay, a 3-fold titration series was performed on a subset of 10 COVID-19 convalescent donors. A 1:2700 plasma dilution was chosen as the optimal dilution to avoid a prozone effect at lower dilutions. **(E)** A decay in %association could be observed from timepoint 1 to timepoint 2 and **(F)** %association at 1:2700 correlated significantly with AUC of the titration curves. Correlations were performed with the non-parametric Spearman test.

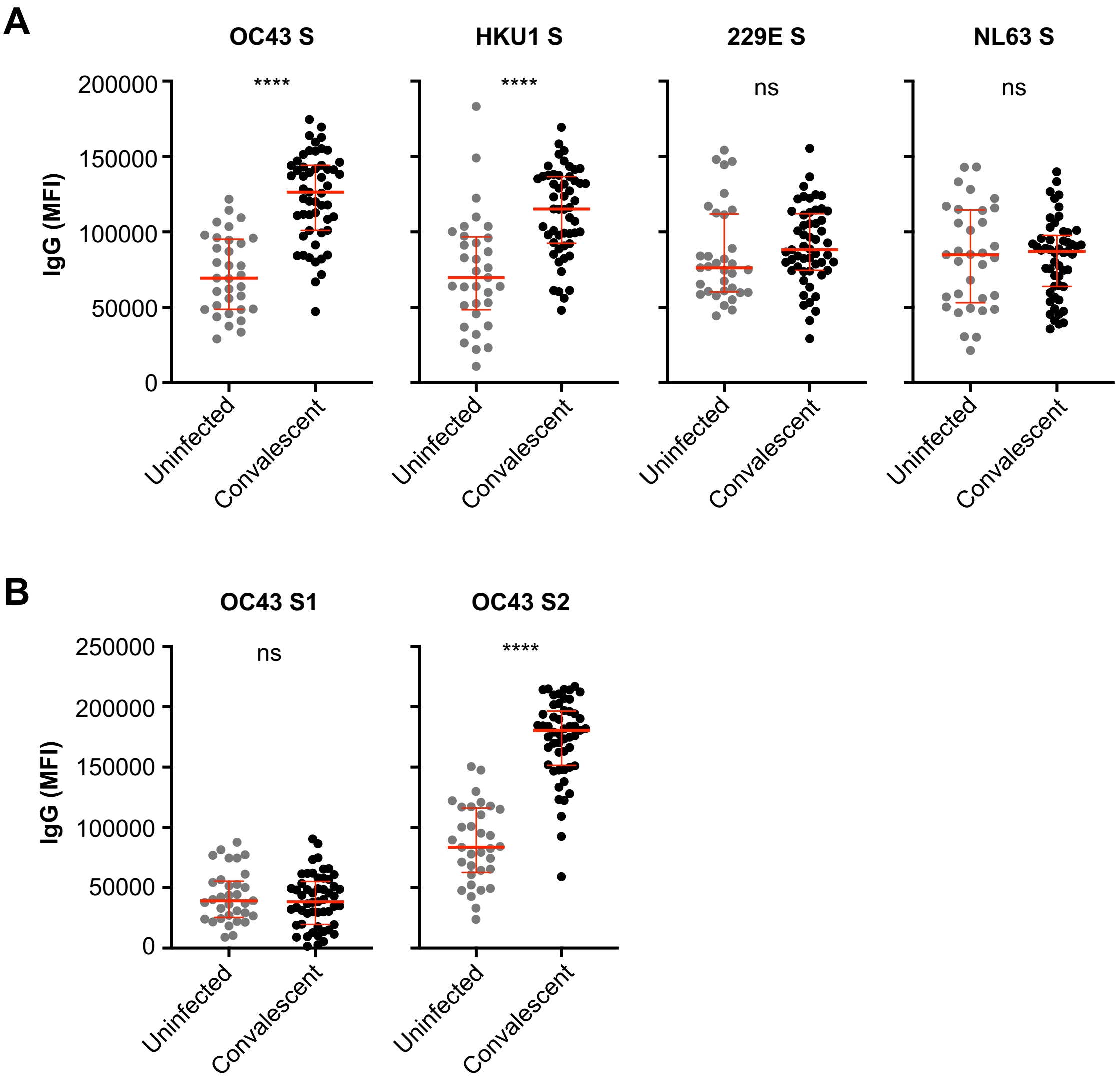


**Fig. S7. The level of IgG antibodies against HCoV S antigens in uninfected controls and COVID-19 convalescent individuals.** Binding of IgG antibodies against **(A)** S from OC43, HKU1, 229E and NL63 and **(B)** S1 and S2 subunits from OC43 in uninfected controls (n=33) and COVID-19 convalescent individuals (n=53) at the first time point. Median and IQR are shown in red lines. Statistical analyses were performed with the Mann-Whitney test (ns, non-significant, ****, *p*<0.0001).

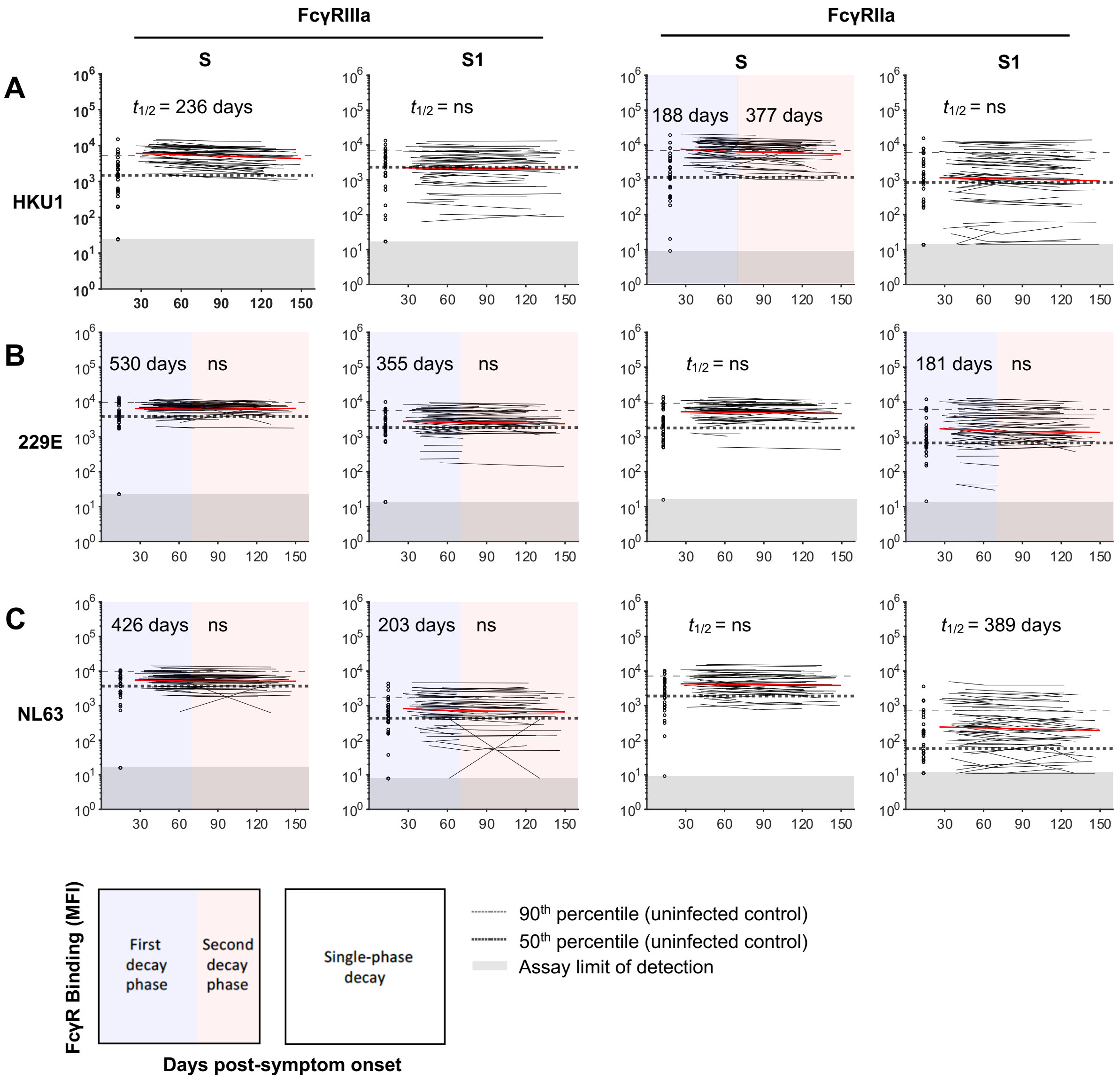


**Fig. S8. Dynamics of dimeric FcγR-binding antibodies against HCoV S antigens (S and S1 subunit) in COVID-19 convalescent individuals.** Kinetics of dimeric FcγRIIIa (F158) and FcγRIIa (R131) binding antibodies against **(A)** HKU1 S and S1, **(B)** 229E S and S1 and **(C)** NL63 S and S1 over time. The best-fit decay slopes (red lines) and estimated half-lives ($t_{1/2}$) are indicated for COVID-19 convalescent individuals. Uninfected controls (n=33) are shown in open circles, with the median and 90% percentile responses presented as thick and thin dashed lines respectively. The limit of detection is shown as the shaded area.

**Supplementary Table 1** Characteristics of the COVID-19 convalescent and uninfected cohorts

| Characteristic | Convalescent cohort (n=53) | Convalescent controls (n=33) | Functional assay cohort* (n=36) | Functional assay controls (n=8) |
|---|---|---|---|---|
| Age, median (IQR) | 55 (49-61) | 53 (27-60) | 56 (49-63) | 58 (45-62) |
| Gender, female (%) | 25 (43.1) | 17 (51.5) | 15 (41.7) | 3 (37.5) |
| Disease severity | | | | |
| Mild (%) | 40 (69) | - | 23 (63.9) | - |
| Moderate (%) | 13 (22.4) | - | 9 (25) | - |
| Severe (%) | 5 (8.6) | - | 4 (11.1) | - |
| Days post-symptom onset | | | | |
| First visit, median (IQR) | 41 (36-48) | - | 41 (38-48) | - |
| Last visit, median (IQR) | 123 (86-135) | - | 131 (121-138) | - |
| Days between first and last visits, median (IQR) | 81 (47-89) | - | 89 (79-95) | - |

*36 subjects from the convalescent cohort with 2 samples at least 60 days apart were chosen for the functional ADCC and ADP assays

**Supplementary Table 2 List of antigens used for multiplex bead array**

| Pathogen | Protein | Vol. coupled / $12.5 \times 10^6$ beads | Source | Cat # | Expression | Tag | Accession # | Amino Acid |
|---|---|---|---|---|---|---|---|---|
| SARS-CoV-2 | S1 subunit | 100μg | Sino Biological | 40591-V08H | HEK293 | His | YP_009724390.1 | Val16-Arg685 |
| SARS-CoV-2 | S2 subunit | 100μg | Acro Biosystems | S2N-C52H5 | HEK293 | His | QHD43416.1 | Ser686-Pro1213 |
| SARS-CoV-2 | Receptor binding domain | 49.7μg | BEI / Florian Krammer | NR-52366 | HEK293 | His | QHD43416 | Arg319-Phe541 |
| SARS-CoV-2 | Trimeric S | 100μg | In-house | | HEK293 | His | YP_009724389.1 | Met—Lys1208 |
| HCoV-229E | S1 subunit | 100μg | Sino Biological | 40601-V08H | HEK293 | His | APT69883.1 | Cys16-Asn536 |
| HCoV-229E | S | 25μg | Sino Biological | 40605-V08B | Insect | His | APT69883.1 | Cys16-Trp1115 |
| HCoV-HKU1 | S1 subunit | 100μg | Sino Biological | 40021-V08H | HEK293 | His | YP_173238.1 | Met1-Arg760 |
| HCoV-HKU1 | S | 25μg | Sino Biological | 40606-V08B | Insect | His | Q0ZME7.1 | Met1-Pro1295 |
| HCoV-NL63 | S1 subunit | 100μg | Sino Biological | 40600-V08H | HEK293 | His | APF29071.1 | Cys19-Val717 |
| HCoV-NL63 | S | 25μg | Sino Biological | 40604-V08B | Insect | His | APF29071.1 | Met1-Pro1296 |

| | | | | | | | | |
|---|---|---|---|---|---|---|---|---|
| HCoV-OC43 | S1 subunit | 100μg | Sino Biological | 40607-V08H1 | HEK293 | His | AVR40344.1 | Met1-Leu794 |
| HCoV-OC43 | S2 subunit | 25μg | Sino Biological | 40607-V08B1 | Insect | His | AVR40344.1 | Ala766-Pro1304 |
| HCoV-OC43 | S | 25μg | Sino Biological | 40607-V08B | Insect | His | AVR40344.1 | Met1-Pro1304 |
| C. Tetani | Tetanus Toxin | 100μg | Sigma-Aldrich | T3194 | | | | |
| SIV | gp120 | 100μg | Sino Biological | 40415-V08H | HEK293 | His | CAA32487.1 | Gln24-Arg531 |
| Influenza A H1N1 (A/Cali/07/2009) | Hemagglutinin | 100μg | Sino Biological | 11085-V08H | HEK293 | His | ACP44189.1 | Met 1-Gln 529 |
